# Chronic loneliness and isolation phenotypes and physical and mental health in older adults

**DOI:** 10.64898/2026.09.08.26362430

**Authors:** Yuteng Ma, Jessica K. Bone, Rosie Mayston, Qian Gao

**Author notes:** **Corresponding author:** Qian Gao.

## Abstract

Chronic social deficits (persistent feelings of being lonely or isolated) have been linked to premature mortality, yet the underlying physical and mental pathways remain unclear. This study used data from 4,090 older adults in the English Longitudinal Study of Ageing, a longitudinal panel study. Using outcome-wide analyses, we assessed how the chronicity of social deficits was associated with 17 health outcomes across physical, mental, cognitive and behavioural health domains. Regression models showed that chronic loneliness was longitudinally associated with increased risks of worse general health, functional capacity, and mental wellbeing, and more unhealthy behaviours. Chronic isolation and onset of isolation were associated with lower quality of life, whereas isolation remission was positively related to enhanced mental wellbeing and cognition compared to no isolation. Findings suggest an association between chronicity of loneliness and adverse health outcomes, highlighting chronic social deficits as an important correlate of healthy ageing.

## Introduction

Loneliness is characterised by subjective feelings of dissatisfaction with the quantity or quality of social relationships^1^. In contrast, social isolation refers to more objective measures of an absence of social connections ^2^. Older people are vulnerable to social deficits (loneliness or social isolation), with research indicating that about one in four British older adults have reported experiencing loneliness or social isolation ^3,4^, and approximately one in five older adults live in chronic loneliness globally ^5^. Loneliness follows a U-shaped distribution across the lifespan, being more prevalent among younger adults and steadily prominent in older adults aged 60 years and above, with its adverse health effects particularly pronounced in older adults ^6–8^. Similarly, social isolation tends to increase as individuals age, with the oldest adults facing a higher risk of living with social isolation compared to those under 70 years ^8,9^. Both loneliness and social isolation adversely impact physical and mental health in later life, increasing the risk of functional difficulties, heart disease, hypertension, anxiety, depression, and dementia ^7,8^. This could result in increased demands for primary care ^10^ and outpatient services ^11,12^. Older people who live with loneliness ^13–15^ or social isolation ^16,17^ have a greater risk of all-cause mortality.

Chronic loneliness and isolation phenotypes present as a persistent, lasting emotional state of feeling lonely or disconnected over an extended period of time ^5,8,18–20^. In contrast, transient loneliness or isolation represents a subacute phenotype, with different health consequences compared to the persistent trait ^21^. Although there is no consensus threshold for the chronic phenotype, studies commonly define it as loneliness or isolation reported consistently across repeated measurements over years or at consecutive time points ^5,8,20^ . As described in the social-ecological model, social relationships are crucial for promoting physical and mental health ^22^, resulting in individuals with chronic loneliness/isolation being particularly vulnerable to poor health. For example, previous research has suggested that chronically lonely groups are likely to be at a greater risk of all-cause mortality than those experiencing occasional loneliness ^8,15,23^.

In theory, compared to transient phenotypes or no loneliness/isolation, chronic phenotypes may accumulate more adverse health impacts through interconnected biological, behavioural, and psychological pathways ^24–26^. Biologically, chronic social deficits may act as a persistent psychological stressor, triggering prolonged allostatic overload ^27–29^ and stress system overactivation ^30^. These dysregulations could lead to elevated inflammatory responses and weaken immune system function ^31,32^. Both chronic loneliness and isolation are robustly associated with systemic chronic inflammation ^32,33^. Specifically, chronic loneliness can potentially upregulate gene transcription within pro-inflammatory control pathways and inflammatory processes ^34,35^. Chronic social deficits have also been associated with imbalances in the autonomic nervous system, epigenetic changes, and neurobiological alterations, all of which may collectively accelerate the biological ageing process and increase vulnerability to chronic diseases ^36^. Consequently, chronic loneliness and isolation may lead to a greater adverse effect on longevity than transient or short-term exposure.

Given the evidence, there are increasing concerns about the impacts of social deficits on public health ^37^. However, previous research often measures social deficits at a single timepoint, with limited attention to the dynamic transitions into and out of loneliness/isolation or long-term exposure to social deficits. Transient loneliness/isolation may have different health impacts compared to chronic phenotypes ^38^. These phenotypes are theoretically distinct ^39^. A recent systematic review highlighted the need for further longitudinal investigations, particularly on chronic loneliness and isolation^5^. Notably, it remains unclear whether experiencing social deficits chronically has cumulative health risks across diverse health domains. Although chronic loneliness and social isolation have been identified as independent risk factors for premature mortality in older age ^8,15,23^, the physical, mental and behavioural pathways underlying the associations between chronic social deficits and premature mortality have yet to be fully explored ^5^. Further research should build on this evidence by testing a range of factors that could represent potential mechanisms. This could help explore the health outcomes of chronic phenotypes and identify potential modifiable mediators of their association with mortality that could be further examined and ultimately intervened on.

Thus, this study used an outcome-wide approach to assess the physical, mental, behavioural, and cognitive health consequences of chronic loneliness and isolation phenotypes and the transitions between loneliness/social isolation states (i.e. onset and remission) over two years. Our research questions were (i) are chronic loneliness and isolation phenotypes and the transitions between loneliness/social isolation states related to poorer health outcomes in older age? (ii) what are the relationships for physical, mental, behavioural, and cognitive health outcomes? (iii) is there a dose-response relationship between chronicity of loneliness/isolation and health outcomes? We hypothesised that chronic and incident loneliness/isolation would be associated with worse health outcomes, whereas remission would be linked to improvements. Chronic phenotypes would be worse than onset, showing a dose-response relationship. This aimed to inform future research to potentially reveal mechanisms through which chronic social deficits could lead to increased mortality.

## Results

We analysed panel data from the English Longitudinal Study of Ageing (ELSA) from 2012 to 2018, examining how chronic and transient social deficits were associated with physical, mental, cognitive, and behavioural health outcomes using regression models. The study included 4,090 participants with a mean age of 66.7 years (SE=0.13, age range= 54-89 years) (Table 1). The sample comprised 2,221 women (54.3%) and was predominantly White (3,971 participants, 97.1%), with 119 participants from other ethnic groups (2.9%). Overall, 2,896 (70.8%) participants were married, and 1,117 participants (27.3%) had only basic education, with 70.1% of the sample being unemployed or retired.

**Table 1.** Characteristics of study samples (n=4,090)

| Characteristics | (Mean, SE)/ n (%) |
| --- | --- |
| Pre-baseline age | 66.7 (0.13) |
| Sex (Female) | 2221 (54.3%) |
| Education |  |
| No qualifications / basic qualifications | 1117 (27.3%) |
| High school /GED [general educational development] (GCSE / O-level / qualification at age 16) | 1117 (27.3%) |
| College (A-levels/higher education/qualification at age 18) | 1067 (26.1%) |
| Postgraduate (Degree / further higher qualification) | 789 (19.3%) |
| Ethnicity |  |
| White | 3971 (97.1%) |
| Other ethnic groups | 119 (2.9%) |
| Pre-baseline marital status |  |
| Yes | 2896 (70.8%) |
| No | 1194 (29.2%) |
| Pre-baseline total (non-pension) wealth | 3.3 (0.02) |
| Pre-baseline employment |  |
| Not working | 2867 (70.1%) |
| Working | 1223 (29.9%) |
| Loneliness |  |
| None | 3141 (76.8%) |
| Transition out of loneliness | 245 (6.0%) |
| Onset of loneliness | 262 (6.4%) |
| Chronic loneliness | 442 (10.8%) |
| Social isolation |  |
| None | 1550 (37.9%) |
| Transition out of isolation | 1399 (34.2%) |
| Onset of isolation | 192 (4.7%) |
| Chronic isolation | 949 (23.2%) |
| Health outcomes at follow-up (wave 9) |  |
| Self-rated health |  |
| Good | 1558 (38.1%) |
| Poor | 2532 (61.9%) |
| Chronic illness | 2434 (59.5%) |
| Chronic pain | 1771 (43.3%) |
| Difficulties with IADLs | 785 (19.2%) |
| Difficulties with ADLs | 769 (18.8%) |
| Mobility limitations | 2131 (52.1%) |
| Gait speed | 3.2 (0.02) |
| Cognitive health | 18.3 (0.07) |
| Depression | 761 (18.6%) |
| Life satisfaction | 3321 (81.2%) |
| Happiness | 3149 (77.0%) |
| Quality of life | 36.9 (0.09) |
| Sleep problems | 1726 (42.2%) |
| Sedentary behaviour | 949 (23.2%) |
| Smoking | 1767 (43.2%) |
| Alcohol consumption |  |
| Less than once a week | 1947 (47.6%) |
| Once or twice a week | 834 (20.4%) |
| 3 or 4 times a week | 536 (13.1%) |
| 5 or more times a week | 773 (18.9%) |
| Leisure engagement | 2.0 (0.02) |

Note. Results weighted and based on 50 imputed datasets. SE: standard error.

We defined chronic phenotypes as loneliness/isolation reported consistently across two consecutive waves. Onset was defined as reporting loneliness or isolation at the first wave only, whereas remission was defined as solely reporting loneliness or isolation at the second wave. Over two years, the prevalence of chronic loneliness (10.8%) was nearly half that of chronic isolation (23.2%). While remission and onset of loneliness were about 6%, the prevalence of isolation remission was 34.2% and 4.7% had onset of isolation. Over half of the sample (59.5%) had chronic illness, and 18.6% had depressive symptoms.

### General health

Compared with people who were not lonely, chronic loneliness was associated with higher odds of chronic illnesses (OR=1.59, 95%CI=1.24-2.04) and chronic pain (OR=1.71, 95%CI=1.33-2.18), as well as lower odds of good self-rated health (OR=0.45, 95%CI=0.34-0.60) two years later (Figure 1 & Supplementary Table 2). Likewise, the onset of loneliness was associated with increased odds of chronic pain (OR=1.39, 95%CI=1.02-1.89), and decreased odds of good self-rated health (OR=0.57, 95%CI=0.41-0.79) than no loneliness (Figure 2 & Supplementary Table 2). Loneliness remission was still associated with lower odds of good self-rated health (OR=0.69, 95%CI=0.49-0.96) (Figure 3 & Supplementary Table 2). There was no evidence that social isolation was associated with any general health outcomes (Figures 1-3 & Supplementary Table 3).

**Figure 1.**
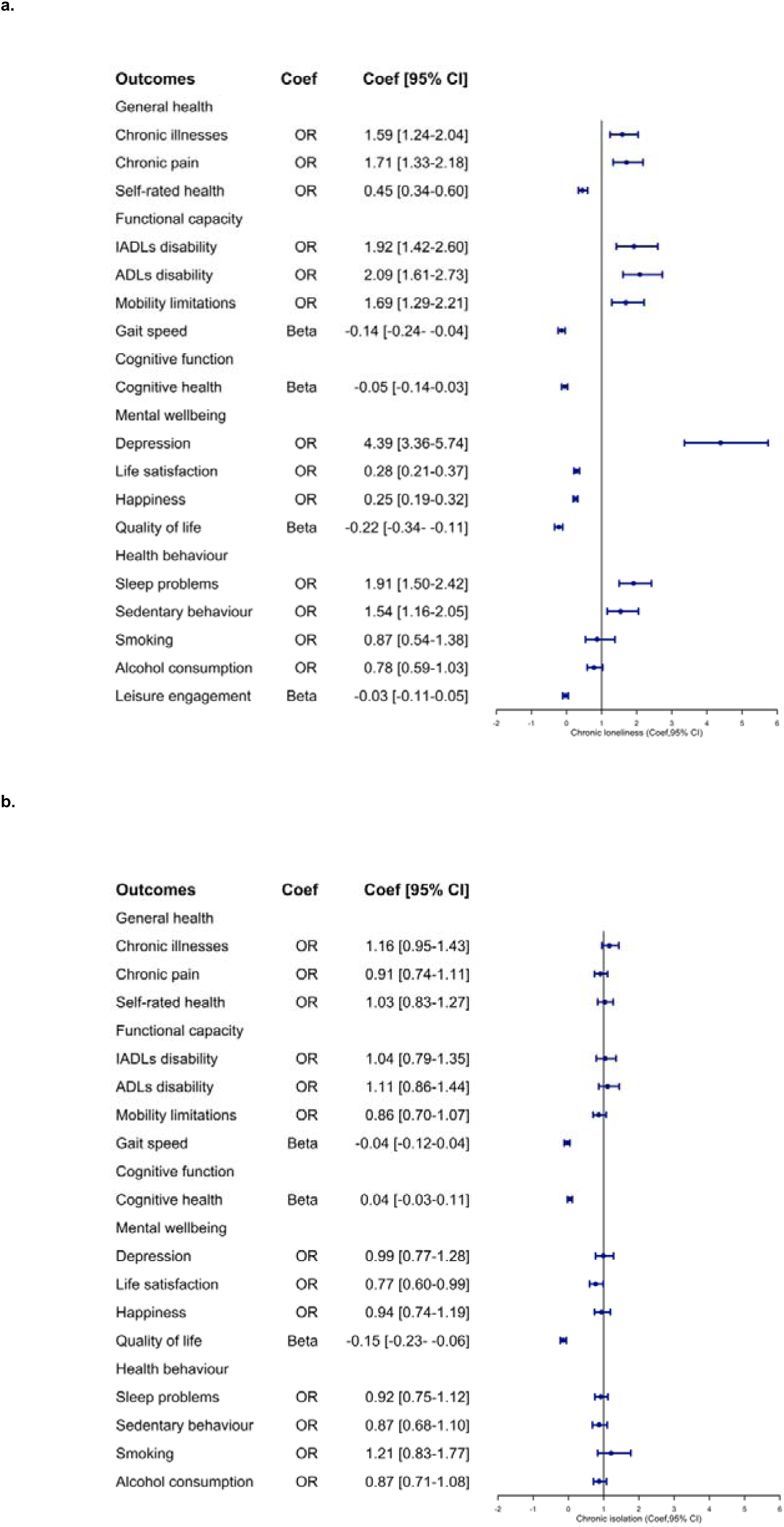
Longitudinal Associations Between Chronic Loneliness and Isolation and Subsequent Physical and Mental Health in Later Life

**Figure 2.**
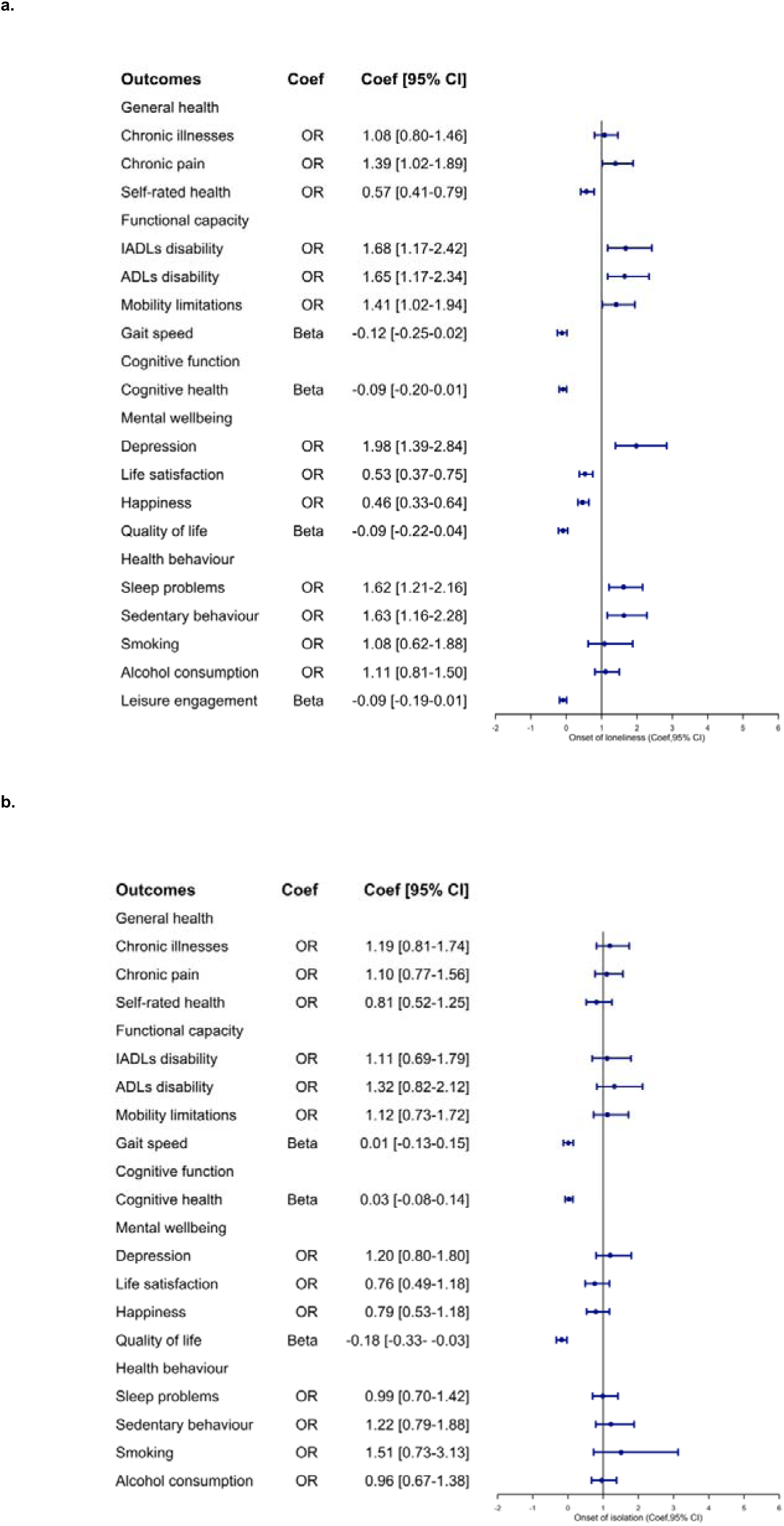
Longitudinal Associations Between Onset of Loneliness and Isolation and Subsequent Physical and Mental Health in Later Life

**Figure 3.**
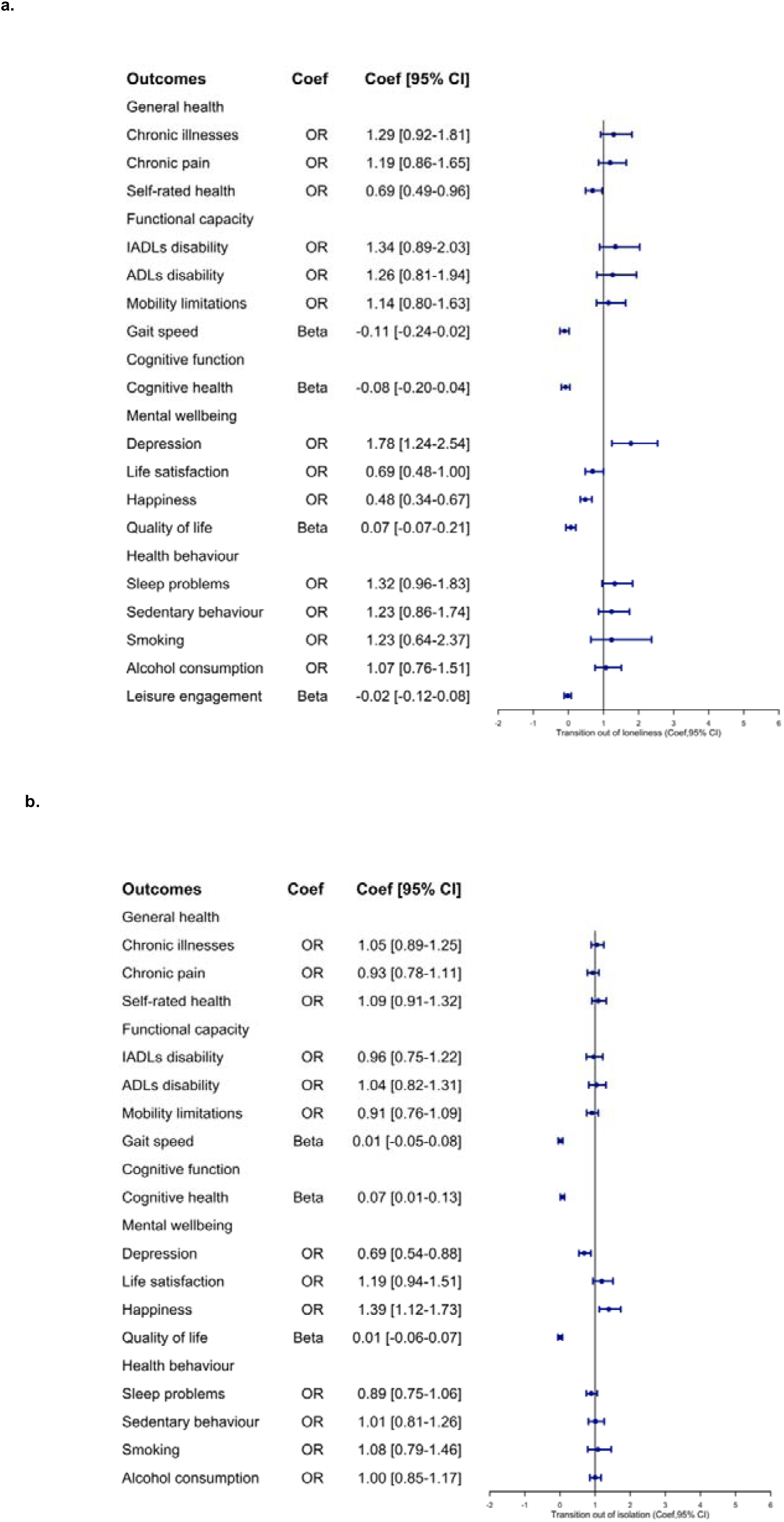
Longitudinal Associations Between Transition out of Loneliness and Isolation and Subsequent Physical and Mental Health in Later Life

### Functional capacity

Independent of confounders, participants with chronic loneliness had increased odds of IADL disability (OR=1.92, 95%CI=1.42-2.60), ADL disability (OR=2.09, 95%CI=1.61-2.73), mobility limitations (OR=1.69, 95%CI=1.29-2.21), and also had slower gait speed (β=-0.14, 95%CI=-0.24 to -0.04) than those who were not lonely. Onset of loneliness was similarly associated with IADL disability (OR=1.68, 95%CI=1.17-2.42), ADL disability (OR=1.65, 95%CI=1.17-2.34), and mobility limitations (OR=1.41, 95%CI=1.02-1.94). No evidence was observed for isolation.

### Cognitive health

There was only evidence for associations between transitioning out of isolation and improved cognitive health (β=0.07, 95%CI=0.01-0.13). Changes in loneliness and other trajectories of isolation were not associated with cognitive health.

### Mental wellbeing

Chronic loneliness was associated with higher odds of probable depression (OR=4.39, 95%CI=3.36-5.74), and with lower odds of life satisfaction (OR=0.28, 95%CI=0.21-0.37) and happiness (OR=0.25, 95%CI=0.19-0.32), and lower QoL (β=-0.22, 95%CI=-0.34 to -0.11). Onset of loneliness was associated with significant but less pronounced higher odds of probable depression (OR=1.98, 95%CI=1.39-2.84), and lower odds of life satisfaction (OR=0.53, 95%CI=0.37-0.75), and happiness (OR=0.46, 95%CI=0.33-0.64). Transitioning out of loneliness was also associated with higher odds of probable depression (OR=1.78, 95%CI=1.24-2.54) and reduced odds of happiness (OR=0.48, 95%CI=0.34-0.67). Chronic isolation was associated with lower life satisfaction (OR=0.77, 95%CI=0.60-0.99). Both chronic isolation (β=-0.15, 95%CI=-0.23 to -0.06) and onset of isolation (β=-0.18, 95%CI=-0.33 to -0.03) were associated with lower QoL. In contrast, transition out of isolation was associated with reduced odds of probable depression (OR=0.69, 95%CI=0.54-0.88) and with increased odds of happiness (OR=1.39, 95%CI=1.12-1.73).

### Health behaviours

Both chronic loneliness and onset of loneliness had higher odds of sedentary behaviour (OR chronic = 1.54, 95%CI=1.16-2.05; OR onset =1.63, 95%CI=1.16-2.28) and sleep problems (OR chronic =1.91, 95%CI=1.50-2.42; OR onset =1.62, 95%CI=1.21-2.16). In contrast, isolation phenotypes were not associated with health behaviour domains.

### Sensitivity analyses

We calculated E-value analyses for associations between chronic loneliness and isolation with health outcomes, with E-values ranging from 1.11 to 8.25, suggesting that findings were moderately robust to unmeasured confounding (Supplementary Table 4). The unadjusted models for longitudinal associations are shown in Supplementary Tables 5-6. We also examined the concurrent associations between chronic social deficits and health outcomes, with findings being generally consistent with longitudinal evidence (Supplementary Tables 7-8).

## Discussion

This study provides a multidimensional assessment of the differential health impacts of chronic and transient loneliness/social isolation phenotypes over time. Our findings indicate that chronic loneliness was longitudinally associated with increased risks of poorer general health (chronic conditions, pain and lower self-rated health), and higher odds of functional disabilities, probable depression, sedentary behaviour and sleep disturbances than no loneliness. Individuals with chronic loneliness were less likely to experience positive psychological wellbeing (including life satisfaction, happiness, and QoL). Both chronic and incident isolation phenotypes were associated with lower QoL, though not with other health domains, including health behaviours. In contrast, transitioning out of isolation was associated with reduced odds of probable depression, greater odds of reporting happiness, and with better cognitive health.

Chronic loneliness is recognised as a risk factor for premature death ^8^, with greater health risks than transient loneliness. Our findings indicate that chronic loneliness potentially accelerate physical, mental and behavioural health risks. This echoes previous research demonstrating that social deficits worsen health via unhealthy behaviours, stress and psychological stressors, and physiological repair mechanisms ^24^. However, no evidence was identified for isolation in our findings, suggesting that the potential health impacts of subjective loneliness and objective social isolation could be explained through different mechanistic pathways ^8^. When examining the transient phenotype, we found that transitioning out of isolation was associated with improved cognitive function, but no evidence emerged for functional improvements after loneliness remission. This differential pattern could be partially explained through the distinct neurobiological mechanisms for loneliness and isolation ^40^. Chronic loneliness may primarily impact health through sustained stress responses ^41^, whereas remission is associated with reduced chronic inflammation that interferes with cognitive health ^32^. Remission may also benefit cognitive health via acute attentional and motivational improvements although these could have a long-lagged effect, warranting further investigations with longer follow-up periods. In contrast, neural changes related to social isolation may be reversible ^42^ , and isolation remission may provide additional cognitive stimulation from new social interaction ^43,44^. Overall, these patterns underscore the conceptual distinction between objective isolation and subjective loneliness, suggesting that future research should explore their distinct mechanisms when designing targeted interventions.

Moreover, our study suggests that chronic loneliness has cumulative negative impacts on both mental and physical health, with potential dose-response relationships between the chronicity of loneliness and adverse health outcomes. These findings suggest that duration and temporal patterns of loneliness may differentially accumulate health hazards through sustained stress-related biological embedding, requiring different intervention priorities. It has previously been suggested that chronic loneliness causes potentially irreversible physiological changes through hypothalamic-pituitary-adrenal (HPA) axis dysregulation and sustained pro-inflammatory responses ^45,46^. Chronic loneliness phenotype may lead to the accumulation of chronic inflammation, which potentially explains the dose-response relationships. Future studies should explore the mechanisms linking chronicity of loneliness to health outcomes, particularly identifying thresholds of experiencing loneliness that significantly alter health risks.

Consistent with previous findings ^7,8^, experiencing chronic loneliness in later life substantially increased risks of developing depressive symptoms and demonstrated negative associations with positive psychological wellbeing (such as life satisfaction, happiness, and QoL). Notably, chronic loneliness and isolation phenotypes demonstrated distinct yet overlapping mental health impacts. Isolation remission had lower risks of experiencing depressive symptoms. However, residual adverse impacts on mental wellbeing (i.e. more depressive symptoms and lower happiness) persisted even after loneliness remission. These persistent “psychological scarring” highlight the potential importance of early identification and intervention for tackling chronic loneliness in future study.

Using a nationally representative sample of England, this study provides insights into the psychosocial health consequences of the chronicity of social deficits. We adjusted for sociodemographic confounders and accounted for pre-baseline outcomes to minimise the risk of adjusting for potential mediators and address reverse causality. Nevertheless, the possibility of omitted variable bias cannot be ruled out. Future studies using causal inference approaches are warranted to further investigate these findings. ELSA sample weights were applied to take account of the complex sample design and sample attrition. However, the study had several limitations. Although well-validated measures were adopted in the survey, the measurement of core indicators primarily relies on self-report methods, potentially introducing recall bias and social desirability bias in assessing the chronicity of social deficits. Depression was measured using the validated CES-D scale, but we did not examine the impact of psychiatric diagnoses on our findings. We limited the sample to those who attended continuous waves of surveys for analytical purposes, which may introduce selection bias (e.g. healthy survival bias). Thus, we weighted the data to mitigate the impact of attrition and sampling biases. But we cannot rule out the possibility of unmeasured confounding and the potential for intermediate confounding during the exposure window. The majority of the ELSA sample were of White ethnicity, which may limit the generalisability of current findings to other ethnicities or cultural settings ^47^. Moreover, the time intervals set for longitudinal assessments were determined by the original survey design, which will not reflect fluctuations in loneliness and isolation between data collection points. Further research is needed to replicate our findings using ecological momentary assessment with different timeframes and in different cultural settings ^48^. Although multidimensional health outcomes were examined, incorporating phenotypic and genotypic biomarkers could better explore the potential mechanisms underlying current findings.

Using an outcome-wide longitudinal approach, this study assessed how chronic phenotypic loneliness and isolation, as well as dynamic transitions between temporary phenotypes, are associated with physical, mental and cognitive health of older adults. We found that chronic loneliness was associated with adverse health outcomes, particularly for mental and physical health. Effect sizes for the onset of loneliness are similar across these domains, yet attenuated relative to the chronic phenotype. In contrast, social isolation was primarily associated with mental health outcomes, and there was some indication that transitioning out of isolation may be linked to improvements in cognitive health. These findings supplement growing evidence that chronic loneliness is linked to later-life health across physical, mental and behavioural health domains, with variation across chronic phenotypes and dynamic transitions. Future research should further elucidate the temporal dynamics underlying these observed patterns and explore whether chronic social deficits might represent a target for intervention to improve healthy ageing.

## Methods

### Ethical approval

This study complies with all relevant ethical regulations. Data were obtained from the English Longitudinal Study of Ageing (ELSA; waves 6-9) through the UK Data Service under the End User Licence. Ethical approval for ELSA Waves 9 and 8 was granted by the South Central— Berkshire Research Ethics Committee (reference numbers 17/SC/0588; 15/SC/0526). ELSA Waves 7 and 6 received ethical approval from the National Research Ethics Service (NRES) Committee South Central—Berkshire (13/SC/0532; 11/SC/0374). All participants provided informed written consent, including consent for the analyses reported in the present study. ELSA participants received tokens of appreciation for their participation and could choose to donate the value of their voucher to charity. These tokens were not intended as financial compensation.

### Data sources

The study used data from the English Longitudinal Study of Ageing (ELSA). ELSA is a longitudinal study including a sample representative of people aged 50 and over living in private households in England. ELSA began in 2002, with follow-up waves conducted every two years ^49^. ELSA data are available from the UK Data Service (https://beta.ukdataservice.ac.uk/datacatalogue/series/series?id=200011). In this study, we used ELSA data from the most recent six-year period, including wave 6 (2012-2013; response rate□= 71.8%-81.8%), wave 7 (2014–2015; response rate□=□78.3%-81.4%), wave 8 (2016–2017; response rate□=□82.4%) and wave 9 (2018–2019; response rate□=□79.5%). To form our final analytical sample, we used waves 7 (2014/2015) and 8 (2016/2017) as our exposure window to measure changes in social deficits over two years. Health outcomes were measured at wave 9 (2018/2019) for longitudinal analyses. Confounders were measured in the wave preceding the start of the exposure window (wave 6; 2012/2013) (Supplementary Figure 1). Of the 7,289 core participants in ELSA at wave 9, we limited our analytical sample to those who participated in all included waves (6-9; n=5,651; 78.2%). In accordance with the primary exposure measures, we then restricted the sample to participants who had complete measures of loneliness and social isolation at waves 7 and 8 (n=4,654). Of these participants, we further limited the sample to those with longitudinal weights provided by ELSA, forming a final sample of 4,090 older adults. The details of sample selection are displayed in Figure 4.

**Figure 4.**
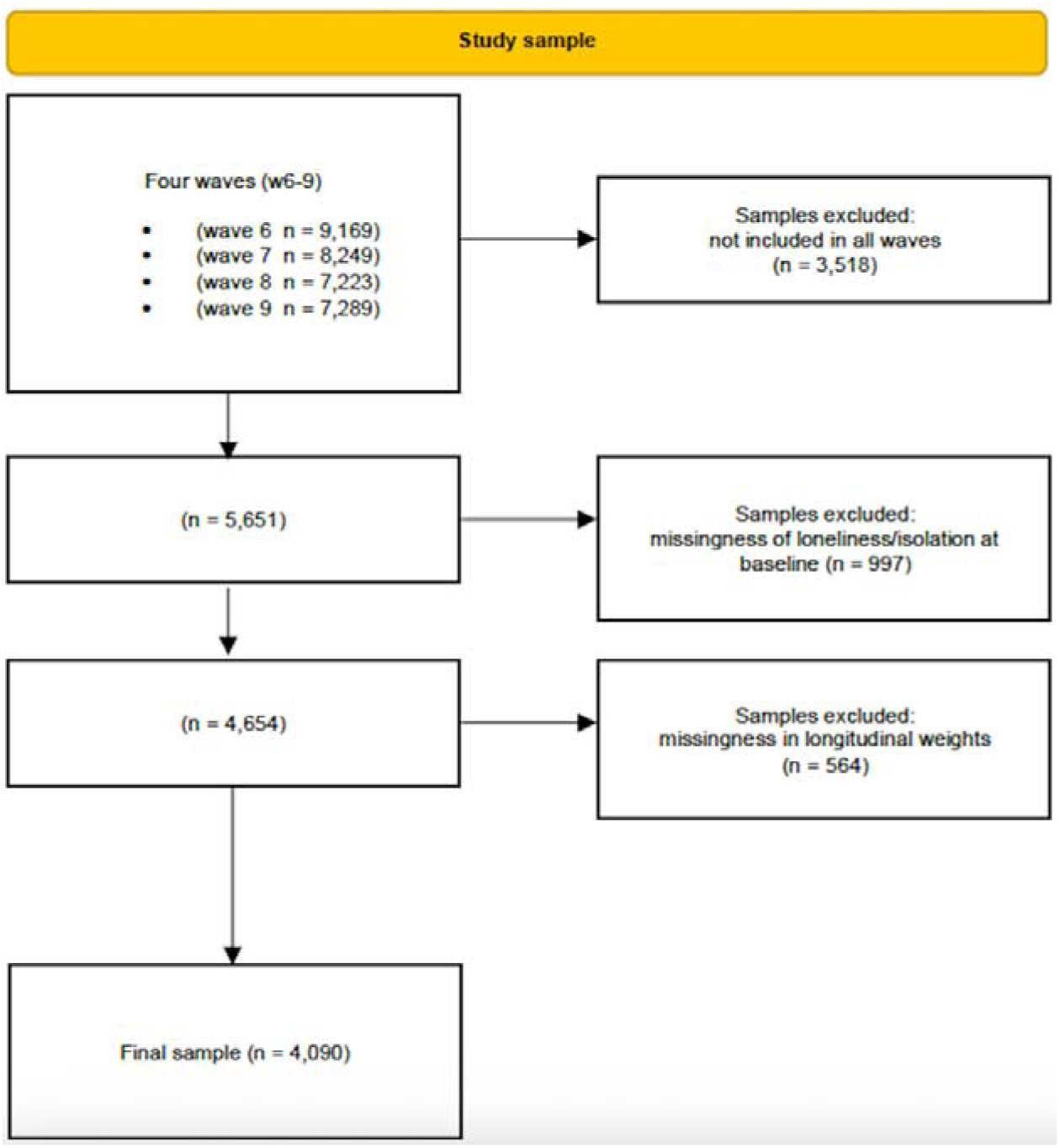
Study sample selection process

### Outcomes

A range of health outcomes were measured at wave 9, within the domains of general health, functional capacity, cognitive function, mental wellbeing and health behaviours.

#### General health

We used a binary indicator of chronic illness (none versus one or more) from a list of common illnesses (including cancer, COPD, arthritis, stroke, diabetes, and angina). Chronic pain was assessed by asking participants if they were often troubled with pain (yes versus no)^50^. Self-rated health status was re-coded as a binary variable (good versus poor).

#### Functional capacity

We generated three binary measures of functional difficulties (yes versus no). First, difficulties with activities of daily living (ADLs) were defined as participants who reported any difficulties with six daily activities (walking across a room; dressing; bathing or showering; eating; getting in or out of bed; and using the toilet) ^51,52^. Second, difficulties with instrumental activities of daily living (IADLs) were defined as any difficulties with managing money; taking medications; grocery shopping; preparing meals; using the telephone; and house/garden work ^53^. Third, mobility limitations were defined as any difficulties with getting up from a chair; climbing one flight of stairs; stooping, kneeling, or crouching; reaching/extending the arms; lifting/carrying weights over 10 lbs; and walking 100 m ^54^. Gait speed was assessed using the validated gait speed test from the Short Physical Performance Battery ^55^. Scores ranged from 0 to 4, with a higher score indicating faster gait speed ^56,57^.

#### Cognitive function

was assessed using a composite indicator of intact mental status and episodic memory, combining scores from several cognitive tasks, including orientation to time ^58^, serial sevens subtraction test, and word-list learning (immediate and delayed recall). Scores ranged from 0 to 29, with higher scores indicating better cognitive function. These tests have been widely used in longitudinal ageing studies, and their reliability and construct validity have been well documented ^59–62^.

#### Mental wellbeing

included both positive and negative health outcomes. Depressive symptoms were measured using the validated eight-item Center for Epidemiologic Studies Depression Scale (CES-D). The scale measured participants’ experience of symptoms in eight domains, with the overall scores ranging from 0 to 8, and a cut-off of ≥3 to indicate probable depression (above-threshold depressive symptom scores) ^63,64^. In accordance with the subjective wellbeing model ^65^, we grouped life satisfaction and happiness under the broader domain of mental wellbeing. Life satisfaction was measured using a 7-point scale (from strongly disagree to strongly agree), and the responses were then recorded as a binary measure (satisfied versus not satisfied). Happiness was measured by asking whether respondents were happy much of the time during the past week (yes versus no). Quality of life (QoL) was captured using the ELSA-adapted version of the CASP-19 measure, a 19-item scale assessing control, autonomy, self-realisation, and pleasure ^66^. Total scores range from 0 to 57, where higher values indicate better QoL ^67^.

#### Health behaviours

included five types of activities. A binary indicator of sedentary behaviour was operationalised as sedentary (less than weekly engagement in mild, moderate or vigorous sports or activities) versus not sedentary (weekly or more frequent engagement) ^68^. Smoking indicated those who smoke versus those who do not smoke. Alcohol consumption included four categories of drinking (less than once a week, once or twice a week, 3 or 4 times a week, and 5 or more times a week). Sleep problems were self-reported as whether respondents felt their sleep was restless during the past week (yes versus no). Finally, leisure engagement (only included in models for loneliness) was measured as participating in volunteering, attending educational or training courses, charity work or associations, social or sports clubs, non-religious community activities, and hobbies, and summarised as overall scores (ranging 0-6), with higher scores indicating higher levels of engagement.

### Exposures

Loneliness was measured using the 3-item UCLA scale ^69^, with a sum score ranging from 3 to 9 and a cut-off score of ≥6 used to indicate loneliness ^70,71^. Social isolation was measured using the five-item Steptoe Social Isolation Index by combining isolation in marital status (unmarried or living alone), less than monthly contacts with children, relatives, or friends, and no social engagement in any groups, clubs or other social organisations, with an overall score ranging from 0 to 5. We applied a cut-off at ≥3 to indicate social isolation ^3,71,72^. We used these measures across waves 7 and 8 to generate two four-category summaries of sustained social deficits over two years: none (not lonely at wave 7 and 8), transition out of loneliness (remission, lonely at wave 7 and not at wave 8), onset of loneliness (not lonely at wave 7 and lonely at wave 8), and chronic loneliness (lonely at wave 7 and 8). We repeated this procedure for social isolation.

### Covariates

We considered a set of pre-baseline covariates including age (a continuous variable), sex (male versus female), ethnicity (White versus other ethnic groups), educational level (no or basic qualifications, GCSE or O-level or qualification at age 16, A-levels or higher education or qualification at age 18, degree or further higher qualification), marital status (married versus not), total net non-pension wealth (quantiles), and employment status (working versus not working) from wave 6.

### Statistical analysis

An outcome-wide approach was applied in the study to understand the associations between chronicity of loneliness and social isolation with a range of subsequent health outcomes, including general health, functional capacity, cognitive function, mental wellbeing, and health behavioural factors. Loneliness and social isolation were tested in separate models. We also ran models separately for each outcome, using logistic regression for binary outcomes, ordered logistic regression for ordinal outcomes, and linear regression for each continuous outcome. Continuous outcomes were standardised. Results are reported as odds ratios (OR) with 95% confidence intervals (CI) for categorical outcomes and beta coefficients (β) with 95%CI for continuous outcomes. All models adjusted for the same set of sociodemographic confounders, including age, sex, ethnicity, education, employment, marital status (only included in models for loneliness), and total net non-pension wealth, and also account for each outcome from pre-baseline (wave 6) to help reduce the risk of adjusting for potential mediators ^73^ and to address reverse causality. Cognitive measures were only available from ELSA wave 7; thus we adjusted for cognitive scores from wave 7 as confounders in modelling. All analyses were conducted on imputed data and weighted using longitudinal weights to account for sample attrition and complex sampling design. The percentage of missingness in each variable ranged from 0% to 25.8% (Supplementary Table 1). All study outcomes, exposures and longitudinal survey weights were included in multiple imputation models, using ordinary least squares (OLS) linear regression, truncated regression, ordered logistic, and logistic regression appropriate to variable types. Datasets were imputed using a multiple imputation by chained equations procedure, and 50 datasets were generated, with estimates combined using Rubin’s rule ^74^. In sensitivity analyses, we examined the concurrent associations between social deficits and health outcomes at wave 8. We also estimated E-values to evaluate the robustness of findings to potential unmeasured confounding ^75,76^. A higher E-value indicates a more robust finding, suggesting that the results are less likely to be attributable to unmeasured confounding. Data analyses were performed using R software version 4.3.2 and Stata 17 (StatCorp LP, Texas, USA).

## Data Availability

ELSA data are available through registration with the UK Data Service (https://beta.ukdataservice.ac.uk/datacatalogue/series/series?id=200011).

## Acknowledgements

The English Longitudinal Study of Ageing is funded by the National Institute on Aging (R01AG017644), and a consortium of UK government departments coordinated by the National Institute for Health and Care Research (NIHR, Ref: 198-1074). The funding bodies had no role in the study design and the collection, analysis, interpretation of data, decision to publish, or preparation of the manuscript. Q.G. is grateful for support from the NIHR Imperial Biomedical Research Centre (BRC). The views expressed are those of the authors and not necessarily those of the affiliated institutions or funding bodies.

## Author Contributions

Y.M. and Q.G. conceived and designed the study. Y.M. acquired and verified the data and conducted the analyses. Y.M. and Q.G. drafted the manuscript. All authors (Y.M., J.K.B., R.M., Q.G.) contributed to the writing and made critical revisions. All authors approved the final manuscript.

## Competing Interests

The authors declare no competing interests.

## Notes

### Competing Interest Statement

The authors have declared no competing interest.

### Author Declarations

Ethical approval for ELSA Waves 9 and 8 was granted by the South Central-Berkshire Research Ethics Committee (reference numbers 17/SC/0588; 15/SC/0526). ELSA Waves 7 and 6 received ethical approval from the National Research Ethics Service (NRES) Committee South Central-Berkshire (13/SC/0532; 11/SC/0374).

